# Benefits of Statins as Adjunct Therapy in the Management of Tuberculosis in Patients at a Referral Site in Ghana

**DOI:** 10.1101/2024.12.18.24319254

**Authors:** Nana Kwaku Bugyei Buabeng, Obed Kwabena Offe Amponsah, Nana Kwame Ayisi-Boateng, Paul Atawuchugi, Kyei Emmanuel Boateng, Aliyu Mohammed, Kwame Ohene Buabeng, Cynthia Amaning Danquah

## Abstract

**Background:** Repurposing medications that have demonstrated efficacy experimentally for drug-resistant tuberculosis (TB), such as statins, provides an opportunity to sidestep the time span and financial resources required to produce new antimicrobials to surmount resistance. However, this has not been extensively explored particularly in sub-Saharan Africa. This study investigated the benefits of statins as adjunct therapy in patients with TB, including those that are drug-resistant.

**Methods:** This was a case control study, involving the review of 3-year medical record of patients with tuberculosis on TB therapy with statins as adjunct (the cases), and those on TB therapy but not on statins at the Agogo Presbyterian Hospital in Ghana. The data obtained was analyzed using STATA version 17. Chi square test was used to determine the association between statin use as adjunct for TB therapy and improved outcomes (cure rate).

**Results:** Statin use in TB therapy as adjunct was associated with an increased incidence of TB cure (p<0.001). The relationship between statin and TB cure was still statistically significant, when age was adjusted for as a confounder in statin exposed individuals, (RR=4.9, p<0.05). Additionally, statins increased the cure rate of TB in the population studied, 6.61 per 100 population, 95% CI, (4.05-10.79) compared to 1.30 per 100 population 95 % CI, (0.65-2.59) in the absence of statins.

**Conclusion:** Statin use in patients on TB therapy was associated with increased cure. This study has affirmed that statins have potential benefits as an adjunct therapy in TB management.

## Introduction

Tuberculosis (TB) claims about 4,000 lives every 24 hours with approximately 1.2 - 1.5 million deaths per year [1]. About 2 billion people were estimated to be infected worldwide [2]. The prevalence of TB in Ghana is about 356 per 100,000 population of bacteriologically confirmed TB raising a big public health concern [3]. High global mortality for tuberculosis can be attributed to the emergence and surge in resistant strains of *Mycobacterium tuberculosis* i.e., multi-drug-resistant TB, extensively-drug-resistant TB (XDR-TB) and in recent times total- drug-resistant TB [4, 5].

The scientific community is challenged to meet up with the demands of increased cases of drug-resistant tuberculosis with limited scope of anti-bacterial agents. The barriers involved in drug development serve as a great limitation to WHO’s end TB strategy as the world fails to catch up with drug-resistant tuberculosis [6]. It may be unsustainable to invest and develop antibiotics only for organisms to develop resistance to their action. It is therefore imperative to consider host-directed therapy where already developed molecules are tested for their ability to intercept the life cycle of TB and then be repurposed for the management of tuberculosis and drug-resistant TB. The repurposing of these drugs would significantly reduce the lead time and cost from bench to bedside [7, 8].

The lipid-lowering pharmaceutical class known as statins is used for both prevention and management of cardiovascular disorders. In early investigations, they have demonstrated potential against MTB infection [9]. Cholesterol reduction and inhibition of isoprenoid intermediates by statins have been shown to reduce *M. tuberculosis* viability in vitro and in vivo [9–11]. A population study by Tahir et al [10]. established that statins may have minimal but statistically significant risk reduction of the incidence of pulmonary tuberculosis.

In sub-Saharan Africa, including Ghana, to the best of our knowledge, there is no available evidence on the use and effectiveness of statins in the management of tuberculosis or drug-resistant strains of tuberculosis. This information is necessary due to the rapid rise in the cases of MDR-TB and XDR-TB in the region compared to other WHO regions [12, 13]. We therefore aimed to assess the benefit of statins as adjunct therapy in the management of tuberculosis through a retrospective study at a TB referral site in Ghana.

## Methods

### Study design and Setting

A retrospective case control study was conducted over a 3-year period (January 2019- December 2021).

The study was conducted at the tuberculosis clinic of the Presbyterian Hospital, Agogo in Ghana. The Presbyterian Hospital is a Quasi-government hospital located in the Asante Akyem South District of Ghana’s Ashanti region. It has a 350-bed capacity and provides specialist services (Dental, infectious disease, medicine, ophthalmology, surgery, obstetrics and gynaecology). The hospital records about 127,000 out-patient attendance annually.

The hospital has a dedicated infectious disease unit, with well-trained physicians, nurses and pharmacists. It is also the main tuberculosis diagnostic centre of the Asante Akyem North District with approximately over 100 active cases of tuberculosis per annum (https://agogopresbyhospital.org/main/profile/).

### Study Population

The sample size was calculated using the Cochran formula [14]. This sampling technique has been well utilized and accepted globally in sample size determination [15].

An expected sample size of 150 was projected for an adequately powered study, we used a conservative estimate of 50%, precision of 5% at 95% confidence interval. Patient records of participants who met the requirements for inclusion were followed from the time the diagnosis of tuberculosis was made, till an endpoint of cure, death or referral. Patients studied were then stratified into individuals on statin therapy and those not on statin therapy.

### Inclusion and Exclusion Criteria

The inclusion criteria for the study were patients diagnosed of tuberculosis from the year 2019 to 2021 and patients with tuberculosis on statin therapy during the study period. Pregnant women and individuals below the age of 25 years were excluded from the study. Pregnant women are unlikely to be on statins due to the concern about the teratogenic effects of statins on fetal development [16].

### Data Collection

Anonymized data were obtained from records of participants who were eligible to be included in the study. Data was obtained from the hospital records. Research assistants were trained on data extraction to ensure accuracy and quality of data. A pilot study was carried out prior to the main study to ensure reliability of data retrieved from the hospital’s electronic management system. A data base in Microsoft Excel format was used as a data collection tool.

Data for this research was assessed for research purposes from September 12, 2022 to January 8, 2023.

### Data Management and Analyses

The data was analyzed using Stata/SE 17.0 Statistical software (Statacorp.4905 Lakeway Drive College Station, Texas 77845, USA). Baseline characteristics were presented as frequency and percentage for categorical variables. Continuous variables were summarized as means and standard deviation for normally distributed variables, and the median and interquartile range for skewed variables. Chi square test was performed to determine the association between statin use in the TB patients on therapy and outcomes. Rate of cure per 100 persons and its 95% confidence intervals were estimated for the respondents. Cox regression analysis was performed to estimate the factors associated with the rate of cure for respondents on statins. A p-value of < 0.05 was considered statistically significant.

### Ethical Considerations

Approval to conduct the study was sought from the research committee of Agogo Presbyterian Hospital and ethical clearance was granted by the KNUST Committee on Human Research, Publications, and Ethics (CHRPE/AP/528/22).

## Results

### Patient baseline characteristics

Data of a total of 170 patients was obtained for the study. The mean age and median age were 43 years respectively, indicating a balance in central tendency. Majority (69.41%) of them were male.

For the population studied, 89.29% were newly diagnosed patients with TB, with 10.71% being cases of relapsed Tuberculosis. Patients with smear negative pulmonary tuberculosis constituted 53.09%, with 37.65% being smear positive. Fifteen (15) patients, constituting 9.26%, had various forms of extrapulmonary tuberculosis. Patients with HIV and TB co-infection were 19.05%.

In terms of the duration of treatment for tuberculosis, 81.73% of patients in this study received standard TB therapy for 6-months, with 18.27% receiving treatment for Tuberculosis for a period greater than 6 months (i.e. 9-24 months). Approximately 24% of patients were on statin therapy. Those who had completed TB therapy constituted 63.82% of patients. However, such patients were not categorized as cured. Those considered cured after treatment were 19.08%. Approximately 5% of patients studied died within the years under review, with 7.89% of patients defaulting treatment as seen in **Table 1**.

**Table 1.** Indicating the baseline characteristics of 170 patients at the Presbyterian Hospital (Agogo) at the TB clinic from 2019-2021.

| Factors | n | % |
| --- | --- | --- |
| Number of TB patients | 170 |  |
| **Age in years, n=165** |  |  |
| Mean (SD) | 43 (19.57) |  |
| Median (IQR) | 43 (32-55) |  |
| **Age category in years, n=165** |  |  |
| 25- 35 | 53 | 32.12 |
| 35-70 | 99 | 60.00 |
| >70 | 13 | 7.88 |
| **Sex** |  |  |
| Male | 118 | 69.41 |
| Female | 52 | 30.59 |
| **Duration of treatment in months, n=104** |  |  |
| 6 | 85 | 81.73 |
| > 6 | 19 | 18.27 |
| **Treatment status, n=168** |  |  |
| New case | 150 | 89.29 |
| Relapse | 18 | 10.71 |
| **Diagnosis type, n=162** |  |  |
| Extrapulmonary | 15 | 9.26 |
| Pulmonary Smear Negative | 86 | 53.09 |
| Pulmonary Smear Positive | 61 | 37.65 |
| **HIV result, n=168** |  |  |
| 279 | 32 | 19.05 |
| 280 | 136 | 80.95 |
| **Statin therapy** |  |  |
| Yes | 33 | 24.81 |
| No | 100 | 75.19 |
| **Outcome, n=152** |  |  |
| Cured | 29 | 19.08 |
| Not cured | 123 | 80.92 |
| **Outcome of treatment, n=152** |  |  |
| Complete | 97 | 63.82 |
| Cured | 29 | 19.08 |
| Dead | 8 | 5.26 |
| Defaulted | 12 | 7.89 |
| Not cured | 2 | 1.32 |
| Referred | 4 | 2.63 |

**SD: Standard Deviation. IQR: Interquartile Range, n:** Number of patients, HIV- Human immunodeficiency virus.

### Patient baseline characteristics

#### Statin Use and Associated Factors

It was established that there was a statistically significant relationship between age and use of statins among the patients.

For patients on statin therapy, 23 (69.70%) were male with 10 (30.30%) females, however, there was no statistical significance difference between those on statins and gender.

Twenty-three patients on statin therapy, received TB treatment in 6 months, with 5 receiving treatment for TB for a period greater than 6 months. There was no statistically significant difference between statin therapy and the duration of TB treatment.

Regarding treatment status of the patients with TB, 87.88% of patients (n=29) on statin therapy were newly diagnosed of Tuberculosis during the study period, 12.12% of patients on statins (n=4) were relapsed. These patients were previously declared cured with a new smear-positive result TB; the p-value showed no statistical significance. There was no significance between statin therapy on the type of TB case and HIV status **(Table 2).**

**Table 2.**
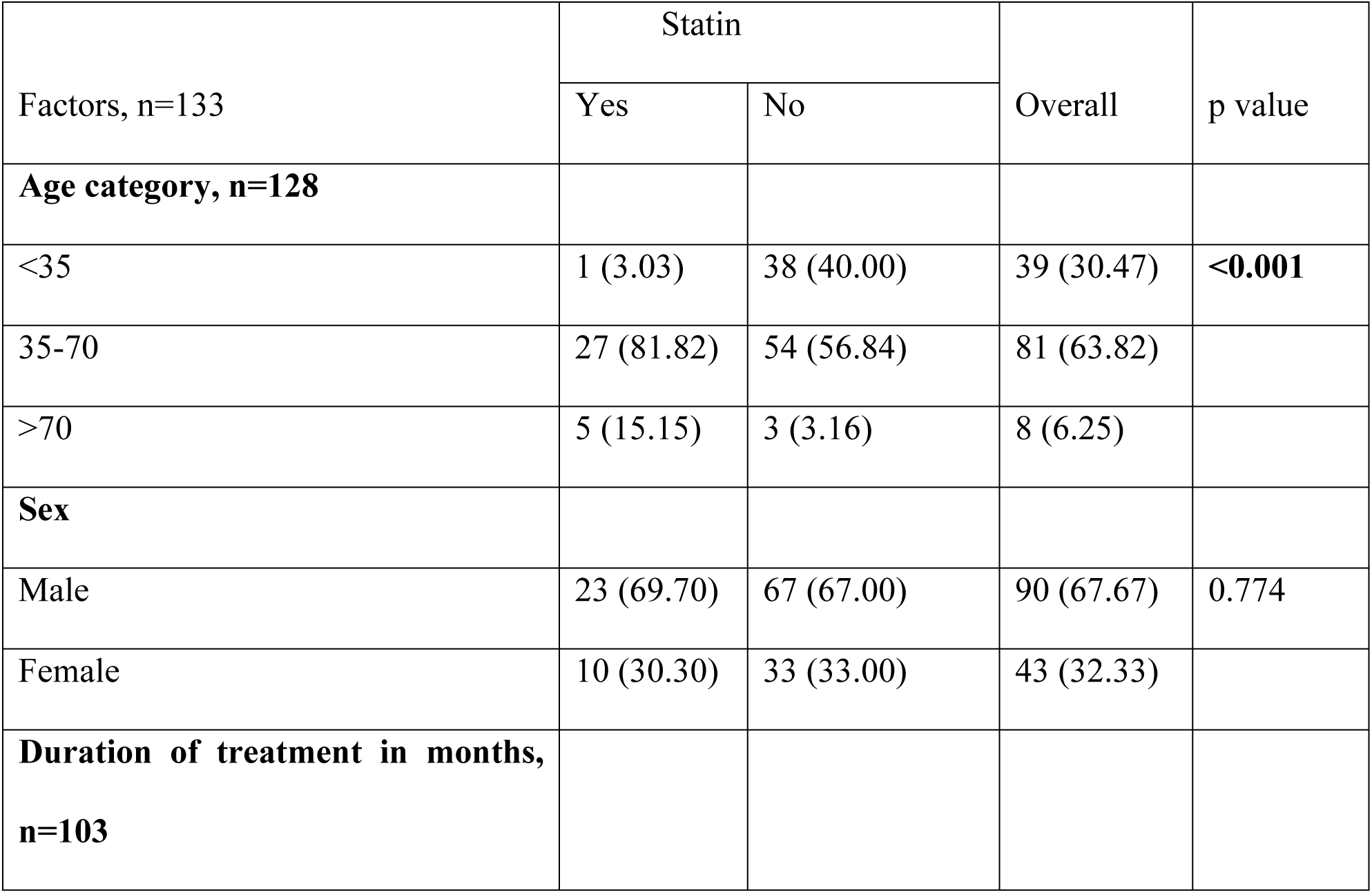
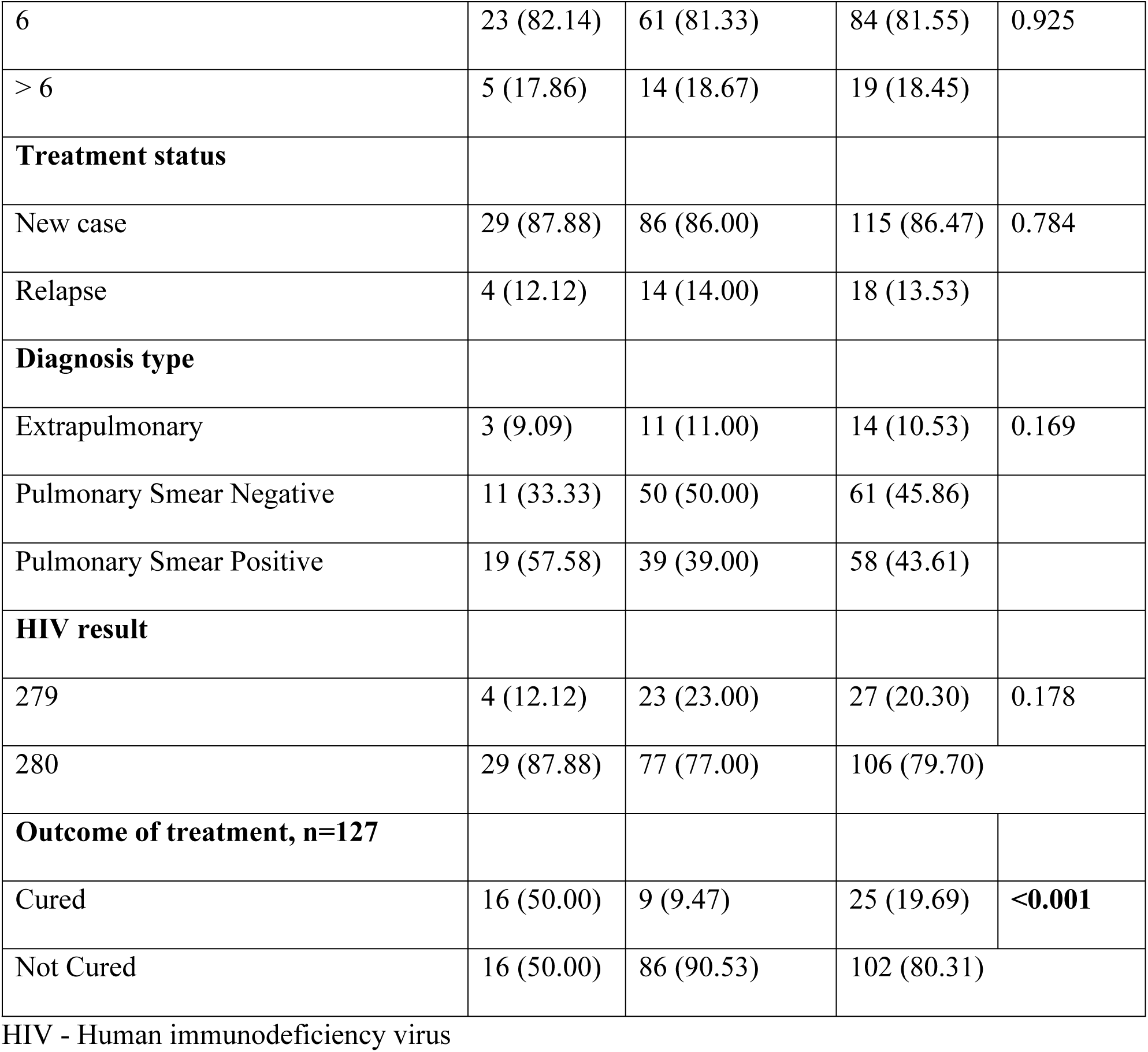
Factors associated with use of statins among patients with TB at the Presbyterian Hospital (Agogo) at the TB clinic from 2019-2021.

Overall, there was a significant association between the use of statins and treatment outcomes for tuberculosis.

### Cure rate of TB in the study population

For every 100 persons who were treated for TB over the study period approximately 3 of them were cured within a period of 9 months required for cure.

Patients with tuberculosis who were on statins had a shorter time (average of 2 months) to cure than those who were not (average of 6 months). Additionally, the rate of cure was higher in the statin group in comparison to the treatment arm not on statin therapy. The cure rates per 100 population was approximately 7 for patients on statin therapy and 1 for individuals not on statin therapy **(Table 3)**.

**Table 3.** The relationship between other associations (Sex and number of co-morbidities) and cure rates for TB patients at the TB Clinic of the Presbyterian Hospital Agogo from 2019-2021.

| Factor | Number Cured | Person-time/months | Cure rate/100 | 95% CI Lower | Upper |
| --- | --- | --- | --- | --- | --- |
| Overall | 24 | 8.65 | 2.77 | 1.86 | 4.14 |
| **Statin** |  |  |  |  |  |
| No | 8 | 6.17 | 1.30 | 0.65 | 2.59 |
| Yes | 16 | 2.42 | 6.61 | 4.05 | 10.79 |
| **Age category** |  |  |  |  |  |
| <35 | 2 | 2.41 | 0.83 | **0.21** | 3.31 |
| 35-70 | 19 | 5.46 | 3.48 | 2.22 | 5.45 |
| >70 | 3 | 0.46 | **6.56** | 2.12 | 20.35 |
| **Sex** |  |  |  |  |  |
| Female | 4 | 2.56 | 1.56 | 0.59 | 4.17 |
| Male | 20 | 6.10 | 3.28 | 2.12 | 5.09 |
| **Co-morbidities** |  |  |  |  |  |
| None | 3 | 1.60 | 1.87 | 0.60 | 5.81 |
| 1 | 4 | 2.73 | 1.47 | 0.55 | 3.91 |
| 2 | 6 | 1.76 | 3.41 | 1.53 | 7.59 |
| 3 | 4 | 1.35 | 2.95 | 1.11 | 7.87 |

Male patients with TB had a higher cure rate than female patients with TB. **Table 2** showed a statistically significant association between the outcome of cure for Tuberculosis and the use of statins with a P value of <0.001 after cox regression analysis.

After adjusting for other covariates, the use of statin was still statistically significant with the cure rate **(Table 4).** The probability of cure on statin therapy was 5 times more likely in the statin exposed group than patients not on statin therapy. Patients above 70 years had a higher probability of cure compared to other age groups.

**Table 4.** Regression analysis indicating the Rate ratio.

| Factor | Unadjusted<br>RR | 95% CI | p<br>value | Adjusted<br>RR | 95% CI | p<br>value |
| --- | --- | --- | --- | --- | --- | --- |
| **Statin** |  |  |  |  |  |  |
| No | 1 |  |  | 1 |  |  |
| Yes | 5.10 | 2.18-11.91 | <0.001 | 4.90 | 1.34-17.92 | **0.016** |
| **Age category** |  |  |  |  |  |  |
| <35 | 1 |  |  | 1 |  |  |
| 35-70 | 4.20 | 0.98-18.03 | 0.054 | 1.65 | 0.31-8.69 | 0.556 |
| >70 | 7.93 | 1.32-47.43 | 0.023 | 8.06 | 1.14-56.95 | 0.036 |
| **Sex** |  |  |  |  |  |  |
| Female | 1 |  |  | 1 |  |  |
| Male | 2.10 | 0.72-6.14 | 0.176 | 1.40 | 0.44-4.35 | 0.566 |
| **Co-morbidities** |  |  |  |  |  |  |
| None | 1 |  |  | 1 |  |  |
| 1 | 0.78 | 0.18-3.50 | 0.748 | 0.75 | 0.15-3.75 | 0.725 |
| 2 | 1.82 | 0.46-7.28 | 0.397 | 0.40 | 0.06-2.54 | 0.333 |
| 3 | 1.58 | 0.35-7.04 | 0.552 | 0.42 | 0.06-2.78 | 0.368 |

P value <0.05 statistical significance

Patients on statin therapy with antitubercular medications had a higher rate of cure than patients without statin therapy **(Figure 1)**. The probability to be cured of TB on statin therapy is two times higher than patients not on statins.

**Figure 1.**
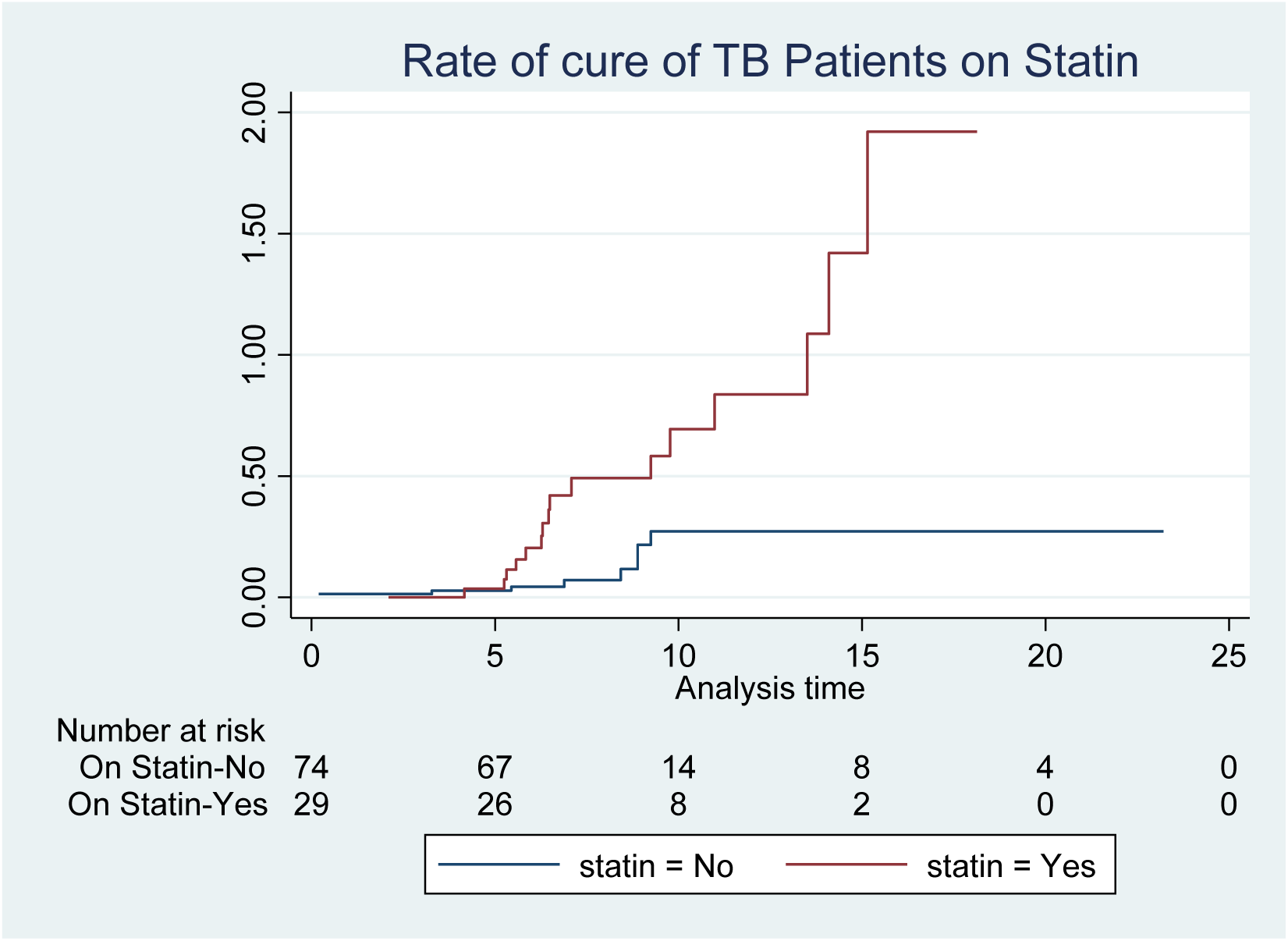
The relationship between cure rate for Tuberculosis for patients on statin therapy against those not on statin therapy at the Presbyterian Hospital (Agogo) at the TB clinic from 2019-2021.

For TB patients on statin therapy, individuals between age 35-70 years had a higher probability of cure than other age categories within the study period **(Figure 2).**

**Figure 2.**
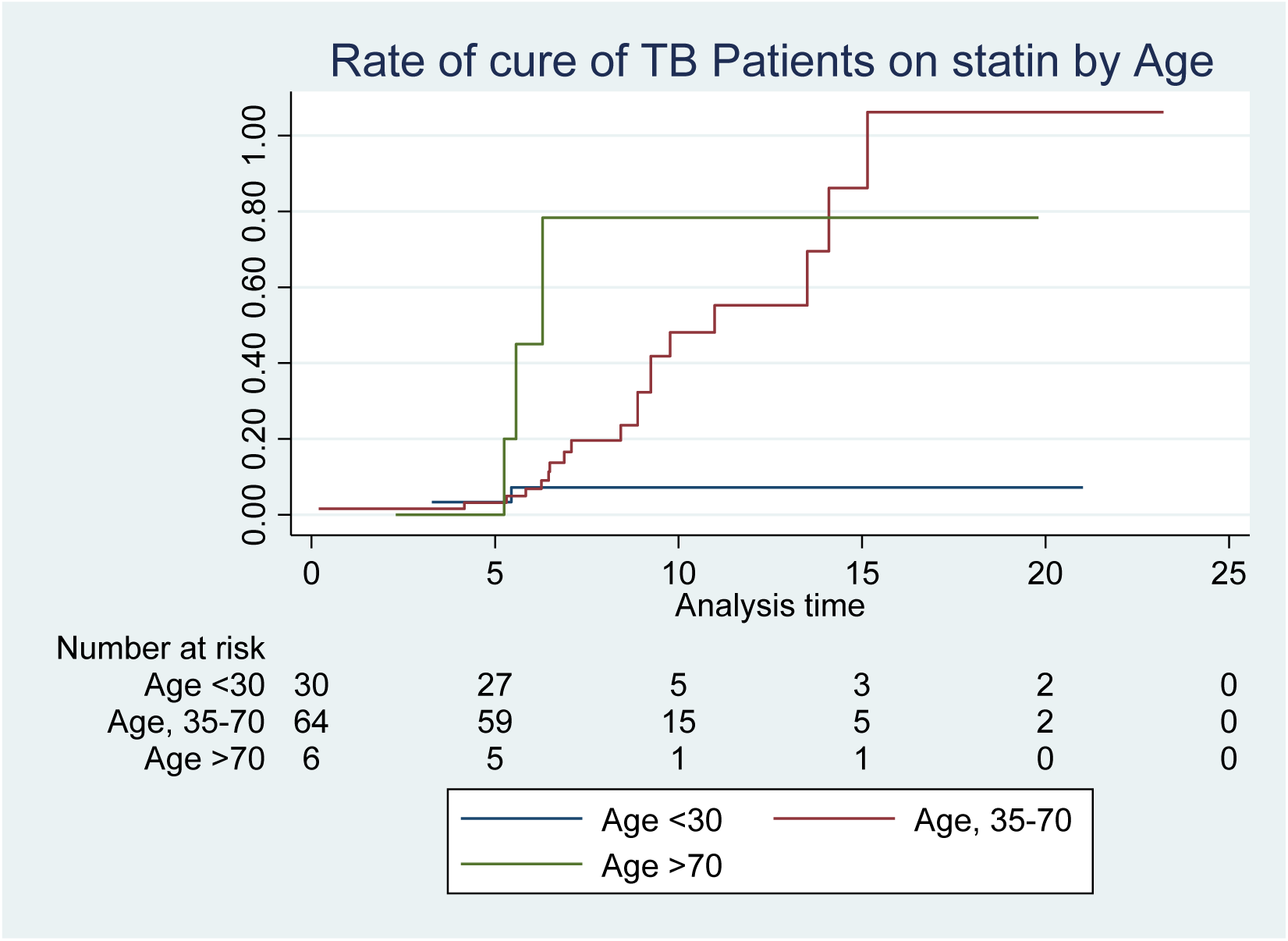
The rate of cure for TB patients on statin therapy after adjusting for age at the Presbyterian Hospital (Agogo) at the TB clinic from 2019-2021.

## Discussion

The study was aimed at assessing the impact of statin therapy in the management of tuberculosis, with a TB referral center as a focal point of the study.

Statin use was statistically related to the rate of cure in the population studied (p<0.001) after therapy. There were two-times increased odds for cure in patients on statin compared to individuals with tuberculosis not on statin therapy. This may provide an array of hope in increasing the rate of cure from tuberculosis in the Ghanaian population. Analysis from this study showed the potential benefit of statin use in the management of tuberculosis which is similar to information from other population studies [17]. A population-based study in Taiwan concluded that statin use in Tuberculosis reduced the risk of pulmonary tuberculosis with an incidence ratio of 0.62 (95% confidence interval (CI), 0.53–0.72), while another study also concluded that risk reduction in tuberculosis was stronger with chronic and long-term statin users, with an adjusted odds ratio of 0.67 (95% CI, 0.59–0.75) [18, 19]. A nested control study also concluded that the risk from fatal pneumonia was reduced in individuals on statin therapy, a nationally representative study by Lai et al [20] established that statin use further reduced the risk of active tuberculosis [21, 22]. An expanded study would however, be beneficial to further establish the effect of statins on the cure from tuberculosis in Ghana and sub-Saharan Africa.

The average time needed to be cured for the patients diagnosed with TB over the study period was 8.65 months. Patients on statins for the duration of the study required a shorter time (2.42 months) to achieve cure compared to individuals not on statin therapy (6.17 months) as seen in Table 3. This may give further credence to the ability of statins to further shorten the duration of TB therapy when used as adjunct therapy for the management of tuberculosis. The long and extended duration for the management of tuberculosis play a major role in contributing to patients defaulting therapy (poor adherence). Ineffective treatment compliance prolongs the infectivity period, increases the risk of recurrence or death from tuberculosis, and can lead to treatment failure and the establishment of drug-resistant tuberculosis [23]. Establishing the cure rate per 100 population, statin therapy was found to have increased the incidence of cure from TB to 6.61 (95% CI (4.05-10.79)) compared to a cure rate 1.30 per 100 population (95% CI, 0.65-2.59) for TB patients not on statin therapy.

Age is a critical confounding factor to the rate of cure, however, after adjusting for age as a confounder, statin use was still significantly associated to the rate of cure from tuberculosis in the population studied. A study by Barreto-Duarte et al [24] confirmed age as a significant risk factor to progression and mortality from tuberculosis in a national therapy outcomes study in Brazil from 2015- 2022. Expanding this study to a larger population may further explore the benefit of statins on increasing the cure rate and duration of TB infection.

The use of statins was related to age, in a statistically significant association. Majority of patients on statin therapy from this study were between the ages of 35-70 years which is similar to age bracket likely to have some compelling indications for statin use. These include patients with a high 10-year atherosclerotic cardiovascular risk (>20%), patients with hyperlipidemia, secondary prevention for patients with a history of ASCVD (stroke, myocardial infarction, peripheral artery disease) usually found from 35-70 years.[22] A 10-year cardiovascular risk assessment from the RODAM study conducted in Ghana, revealed that individuals aged 40- 70 years had an elevated cardiovascular risk [18]. It is therefore not surprising that individuals in this age bracket are likely to be on statin therapy as seen in this study. Individuals during the study period were on statins for the management of dyslipidemia, as primary and secondary prevention of atherosclerotic cardiovascular conditions including stroke, myocardial infarction, peripheral artery disease among others.

Individuals older than 70 years on statin were noticed to have a higher cure rate per 100 population 6.56 (95% CI (2.12-20.3)). This finding was inconsistent with other age-related studies on tuberculosis. In these studies; older age groups had more advanced disease at the time of diagnosis, and a higher proportion had comorbid illnesses [25, 26]. A larger study may provide more information on the possible protective effect of statins in the management of tuberculosis in the elderly, to firmly establish its protective effect.

In this study, approximately 69% of the patients with TB were males which correlates with the reported higher prevalence of TB in Ghanaian males. This may be attributed to increased exposure to social risk factors of TB (poor sanitation, smoking, alcoholism) [27]. The median age for the patients was 43 years. Attributable reasons may be the health seeking behavior of the people in the study area. Previous studies at the Agogo Presbyterian Hospital reveal that young individuals in the surrounding districts are more likely to seek health care than the elderly [28]. The median age may also increase the risk of ongoing transmission as relatively young people are likely to engage in social activities that may increase spread of this airborne disease.

Majority of diagnosed cases of tuberculosis were smear-negative, in consonance with that seen in other population-based TB studies where >50% of the cases were diagnosed as smear-negative tuberculosis [29, 30].

Human Immunodeficiency virus remains a major risk factor for tuberculosis and is associated with an increased risk of mortality [31]. Patients living with HIV, the commonest co-infection, constituted about a fifth of the participants. This is likely due to the immunocompromised state of HIV patients which increases the risk of acquiring primary tuberculosis [32].

For the period under study, there were no diagnosed cases of multi-drug resistant tuberculosis, in consonance with a previous study in Ghana, where it was determined that a reduction in MDR-TB cases can be attributed to a number of factors; the incorporation of BCG vaccination in the immunization schedule and improved detection of Tuberculosis in communities through improved public surveillance systems [33].

## Strengths and limitations

This study is - to the best of our knowledge - the first to analyze the use of statins in a population of TB patients in Ghana. Secondly, the study may provide a basis for public health interventions in improving outcomes in TB patients who may require statins for various atherosclerotic cardiovascular conditions if this study is expanded. Again, the study was conducted and reported in accordance with the Strengthening the Reporting of Observational Studies in Epidemiology (STROBE) guidelines statement [34]. A limitation to this study includes the fact that the data was obtained from one TB referral site and thus the findings may not be generalizable. A broader multi-center study may be helpful to enhance the generalizability of findings.

## Conclusion

Statin use was associated with increased cure rate from tuberculosis management at the study site. The outcome of this study has affirmed the potential benefit of statin as adjunct therapy in the management of TB, including drug and multi-drug-resistant TB. This calls for more robust assessments, including multi-center trials for stronger evidence to ensure effective TB therapy with appropriate national and global narrative of the role of statins in TB management.

## Data Availability

All relevant data are within the manuscript and its supporting information files.

## Acknowledgements

We acknowledge the support of the management of the Presbyterian Hospital, Agogo- Asante Akyem for granting us access to the Hospital’s data base for this study. We also acknowledge the contribution of the I.T and the infectious disease unit of the Presbyterian Hospital as well as members of the Drug Discovery team of the Department of Pharmacology, Kwame Nkrumah University of Science and Technology, Kumasi, Ghana.

## Author Contributions

**Conceptualization-** Nana Kwaku Bugyei Buabeng, Kwame Ohene Buabeng, Cynthia Amaning Danquah

**Data Curation -** Nana Kwaku Bugyei Buabeng, Nana Kwame Ayisi Boateng

**Formal Analysis -** Nana Kwaku Bugyei Buabeng, Kyei Emmanuel Boateng

**Investigation -** Nana Kwaku Bugyei Buabeng

**Methodology- Nana** Kwaku Bugyei Buabeng, Cynthia Amaning Danquah, Aliyu Mohammed

**Project Administration-** Nana Kwaku Bugyei Buabeng, Cynthia Amaning Danquah

**Resources -** Nana Kwaku Bugyei Buabeng, Cynthia Amaning Danquah

**Software -** Nana Kwaku Bugyei Buabeng, Obed Kwabena Offe Amponsah **Supervision -** Cynthia Amaning Danquah, Nana Kwame Ayisi Boateng

**Validation -** Paul Atawuchugi, Nana Kwame Ayisi Boateng, Kwame Ohene Buabeng, Cynthia Amaning Danquah

**Visualization-** Kyei Emmanuel Boateng, Paul Atawuchugi

**Writing – Original Draft Preparation -** Nana Kwaku Bugyei Buabeng, Obed Kwabena Offe Amponsah

**Writing – Review & Editing -** Nana Kwaku Bugyei Buabeng, Kwame Ohene Buabeng, Cynthia Amaning Danquah, Nana Kwame Ayisi Boateng, Kyei Emmanuel Boateng, Aliyu Mohammed, Obed Kwabena Offe Amponsah, Paul Atawuchugi

**Funding:** This research received no specific grant from any funding agency in the public, commercial, or not-for-profit sectors.

**Informed Consent Statement:** As this was a record review study with no patient identifiers, the issue of informed patient consent did not apply.

**Data Availability Statement:** Reasonable requests to access these data may be sent to the corresponding author.

**Transparency declarations:** None to declare

